# Automated Deep Learning Segmentation of Cardiac Inflammatory FDG PET

**DOI:** 10.1101/2024.01.31.24302113

**Authors:** Alexis Poitrasson-Rivière, Michael D. Vanderver, Tomoe Hagio, Liliana Arida-Moody, Jonathan B. Moody, Jennifer M. Renaud, Edward P. Ficaro, Venkatesh L. Murthy

**Author notes:** Address for correspondence: Alexis Poitrasson-Rivière, PhD, INVIA Medical Imaging Solutions, 3025 Boardwalk Dr., Suite 200, Ann Arbor, MI, 48108. Equal Contributions as co-senior authors (Edward P. Ficaro, Venkatesh L. Murthy).

## Abstract

**Background:** Fluorodeoxyglucose positron emission tomography (FDG PET) with glycolytic metabolism suppression plays a pivotal role in diagnosing cardiac sarcoidosis. Reorientation of images to match perfusion datasets is critical and myocardial segmentation enables consistent image scaling and quantification. However, both are challenging and labor intensive. We developed a 3D U-Net deep learning (DL) algorithm for automated myocardial segmentation in cardiac sarcoidosis FDG PET.

**Methods:** The DL model was trained on 316 patients’ FDG PET scans, and left ventricular contours derived from perfusion datasets. Qualitative analysis of clinical readability was performed to compare DL segmentation with the current automated method on a 50-patient test subset. Additionally, left ventricle displacement and angulation, as well as SUVmax sampling were compared to inter-user reproducibility results.

**Results:** DL segmentation enhanced readability scores in over 90% of cases compared to the standard segmentation currently used in the software. DL segmentation performed similarly to a trained technologist, surpassing standard segmentation for left ventricle displacement and angulation, as well as correlation of SUVmax.

**Conclusion:** The DL-based automated segmentation tool presents a marked improvement in the processing of cardiac sarcoidosis FDG PET, promising enhanced clinical workflow. This tool holds significant potential for accelerating clinical practice and improving consistency and quality. Further research with varied datasets is warranted to broaden its applicability.

## Introduction

Cardiac sarcoidosis results from the infiltration of inflammatory cells into the myocardium, leading to a range of serious cardiac complications including arrhythmia, heart failure, and death [1–3]. Diagnosis of cardiac sarcoidosis may lead to invasive and complex treatments including placement of implantable cardioverter-defibrillators and immunosuppressive medications. Consequently, accurate diagnosis is of utmost importance. Furthermore, serial assessment of inflammatory activity is often helpful to guide management of immunosuppression [4]. FDG PET with metabolic suppression of myocardial glycolytic metabolism [5–7] is critical for the diagnosis and management of cardiac sarcoidosis. In addition, similar types of scans are now an important part of the diagnosis of prosthetic valve endocarditis in both European and American guidelines [8, 9].

Correlation of FDG uptake to perfusion findings is critical for accurate interpretation of cardiac sarcoidosis PET studies [10]. Ideally, this is done in cardiac planes: short axis, vertical long axis, and horizontal long axis. Segmentation of the myocardium allows for scaling of these images to optimize image dynamic range. However, standard cardiac segmentation algorithms developed for myocardial perfusion imaging (MPI) do not perform well for cardiac FDG PET scans performed with suppression of myocardial glycolytic metabolism. This is due to cardiac perfusion studies having mostly uniform distribution throughout the myocardium, whereas FDG PET scans with myocardial suppression have highly variable distributions of activity. Normal studies have no uptake in the myocardium while abnormal studies typically present hot spots of activity in varied locations. Manual segmentation and reorientation are time consuming and inconsistently used in many laboratories. Improper or suboptimal myocardial segmentation could lead to diagnostic errors if incorrect image scaling is applied to FDG PET scan images. Finally, approaches that copy segmentation contours from perfusion to FDG PET scans still require manual intervention and are not possible when perfusion images are not routinely acquired (i.e., for endocarditis FDG PET).

Deep learning (DL) approaches to image segmentations have been increasingly used in the medical imaging field, successfully performing various tedious segmentation tasks [11, 12]. Consequently, we sought to develop a fully automated myocardial segmentation algorithm for cardiac FDG PET performed with suppression of myocardial glycolytic metabolism using a deep convolutional neural network.

## Methods

### Study Population

316 non-consecutive patients with cardiac FDG PET scans performed with a myocardial FDG suppression protocol [5, 13], coupled with rest ^82^Rb perfusion PET scans, at the University of Michigan between June 2015 and June 2018, along with their repeat studies ranging from May 2012 to May 2019, were processed and reviewed in Corridor4DM (INVIA Medical Imaging Solutions, Ann Arbor, MI, USA). There were no exclusion criteria. Contours of the left ventricular myocardium created on perfusion datasets [14] were manually transferred to inflammatory datasets using image fusion by two users (imaging expert and trained technologist). Contours were subsequently converted to binary masks in transverse orientation. The transverse FDG PET volumes and binary masks were separately used as input and label for the supervised DL modeling. The 316 patients were divided into training, validation and testing sets of 189 (306), 63 (112), and 64 (106) patients (studies) respectively for our DL modeling effort. Additionally, a final test population of 50 patients was randomly selected from patients with an inflammatory FDG scan performed in 2019, taking care to exclude any individuals who also had prior scans in the training population. In the case of patients with multiple scans performed in 2019, only the latest scan was selected. The final test population was processed by both users to obtain inter-user variability.

### DL Algorithm

The DL architecture applied in this work is a 3D U-Net model, which has been used previously for image segmentation tasks. The U-Net architecture was originally designed for 2D image segmentation but is extended here to handle 3D volumetric data [15, 16]. The model consists of an encoder-decoder structure. The encoder captures the high-level features and context from the input 3D volume (128×128×80) while the decoder uses this information to generate a segmented output. The encoder uses convolutional layers for feature extraction and pooling layers to downsample the spatial dimensions of the data, reducing computational complexity. In the decoder, upsampling layers are employed to increase the spatial dimensions of the data. Skip connections directly link the corresponding layers in the encoder and decoder, allowing for the preservation of fine-grained details during the upsampling. The final layer consists of a convolutional layer with a sigmoid activation function. This produces the segmentation map, matching the dimensions of the input 3D volume. The model was trained with a binary cross entropy loss function and the Adam optimizer, with a fixed learning rate of 0.001, for 500 epochs. Performance improvement through epochs was also tallied using the dice coefficient, a measure of similarity commonly used for segmentation, of the binary contour masks and the resulting segmentation maps.

### Qualitative and Quantitative Result Analysis

Our final test population was processed by two users (imaging expert and trained technologist) as well as run through the DL segmentation algorithm and the standard surface generator used for perfusion image segmentation provided in Corridor4DM. Segmented images normalized to peak uptake within contours from the DL segmentation algorithm and the standard segmentation algorithm were reviewed by an expert physician reader (100 images). Each image was scored on a scale of 1 to 4 (1:Poor, 2:Fair, 3:Good, 4:Excellent) for quality of: left ventricular centering, left ventricular orientation, scaling/normalization of image intensity, and overall readability. In addition, we compared the centering, orientations, and SUVmax produced by the DL and standard segmentation methods to the inter-user variability with manual contour transfer.

## Results

The demographics of the training, validation, and test subsets as well as our final test population are shown in Table 1.

**Table 1.** Demographics of the 2 patient populations.

|  | *DL Modeling Population* |  |  | *Final Analysis Population* |
| --- | --- | --- | --- | --- |
|  | *Training* | *Validation* | *Testing* |  |
| *N* | 189 | 63 | 64 | 50 |
| *Male* | 128 (68%) | 29 (46%) | 36 (56%) | 32 (64%) |
| *Age (years)* | 57.0 ± 11.4 | 57.7 ± 11.5 | 53.5 ± 13.2 | 60.9 ± 11.4 |
| *Race (AA, C, A/P)* | 46 (24%),<br>134 (71%),<br>1 (1%) | 16 (25%),<br>45 (71%),<br>0 (0%) | 19 (27%),<br>45 (70%),<br>0 (0%) | 11 (22%), 35 (70%), 1 (2%) |
| *BMI (kg/m<sup>2</sup>)* | 31.7 ± 6.7 | 32.0 ± 6.7 | 32.0 ± 7.3 | 30.2 ± 6.4 |
| *Hypertension* | 107 (57%) | 29 (47%) | 36 (56%) | 37 (74%) |
| *Hypercholesterolemia* | 91 (48%) | 27 (44%) | 30 (47%) | 24 (48%) |
| *Diabetes* | 42 (22%) | 12 (19%) | 19 (30%) | 15 (30%) |
| *Smoking* | 28 (15%) | 8 (13%) | 13 (20%) | 7 (14%) |

AA: African American
C: Caucasian
A/P: Asian/Pacific Islander
12 patients were missing race information

### Concordance of Segmented Contours

Figure 1 demonstrates convergence of segmentation with maximization of the Dice coefficient in the validation subset. The model was verified to yield similar performance with the DL modeling test dataset, yielding a dice coefficient of 0.622.

**Figure 1.**
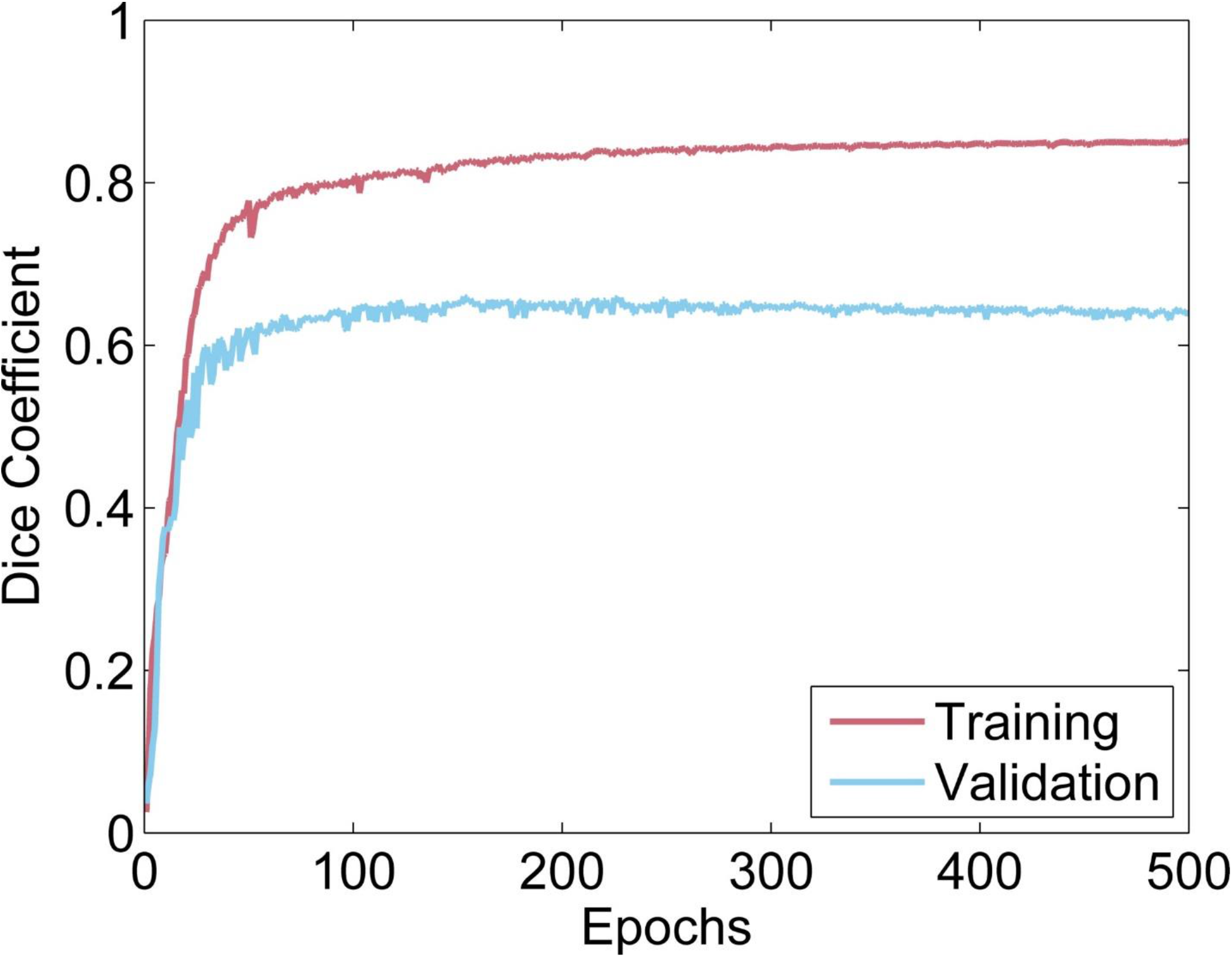
Evolution of the dice coefficient throughout the epochs for the training and validation datasets

### Qualitative Analysis of Automatic Segmentation

To assess the adequacy of DL segmentation for clinical interpretation, the algorithm was compared to the standard segmentation developed and optimized for perfusion images [14]. Blinded physician scores for a random mixture of DL and standard segmentations are shown in Figure 2. Generally, standard segmentation fared poorly, with more than 50% of studies rated as fair to poor. The DL algorithm consistently improved centering, alignment, normalization, and overall readability of studies, yielding scores of good to excellent in more than 90% of studies.

**Figure 2.**
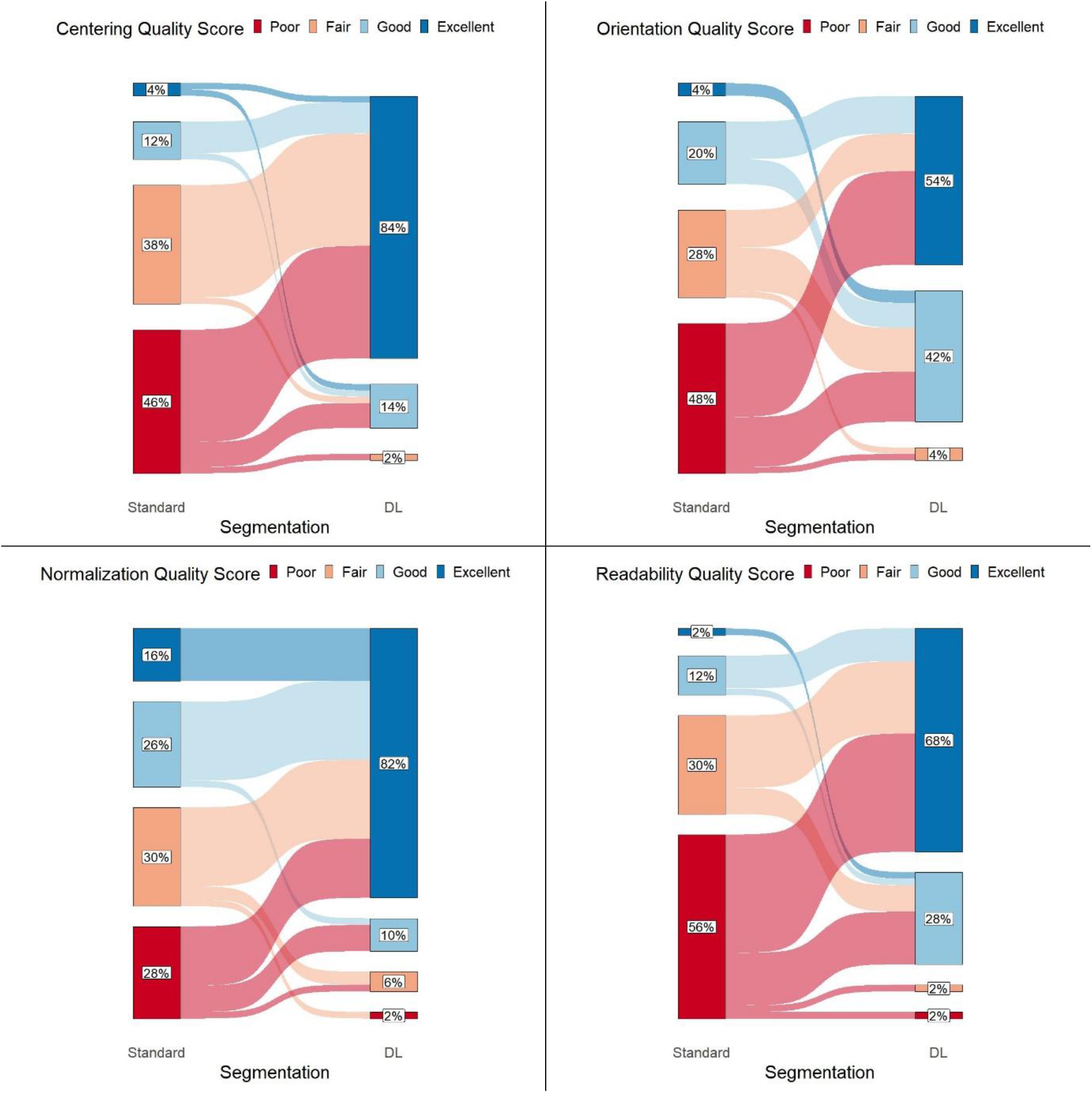
Evolution of quality scores with the introduction of DL segmentation

Only 3 (6%) studies were rated worse with DL segmentation than with standard segmentation: 2 studies decreased from excellent to good for combinations of centering, orientation, and overall readability, and 1 study decreased from fair to poor in normalization (notably being scored as poor in overall readability for both the DL and standard segmentation). Importantly, no ratings decreased from excellent/good to fair/poor. If successful segmentation is defined as having all quality scores greater than 2 (good and above), DL segmentation improves the success rate to 90% from 12% for standard segmentation (p<0.0001). If successful segmentation is defined as having all quality scores greater than 1 (fair and above), DL segmentation improves success to 98%, compared with 32% for standard segmentation (p<0.0001).

### Inter-User Processing Reproducibility

Both standard and DL segmentations were compared to the reproducibility of manual processing between a trained technologist and an imaging expert. Overall, displacement of the left ventricle (LV) center for the DL segmentation compared to the imaging expert was similar to that for the trained technologist and was markedly smaller than for the standard segmentation algorithm (Figure 3a, p < 0.0001). Similar results were observed for angulation in Figure 3b.

**Figure 3.**
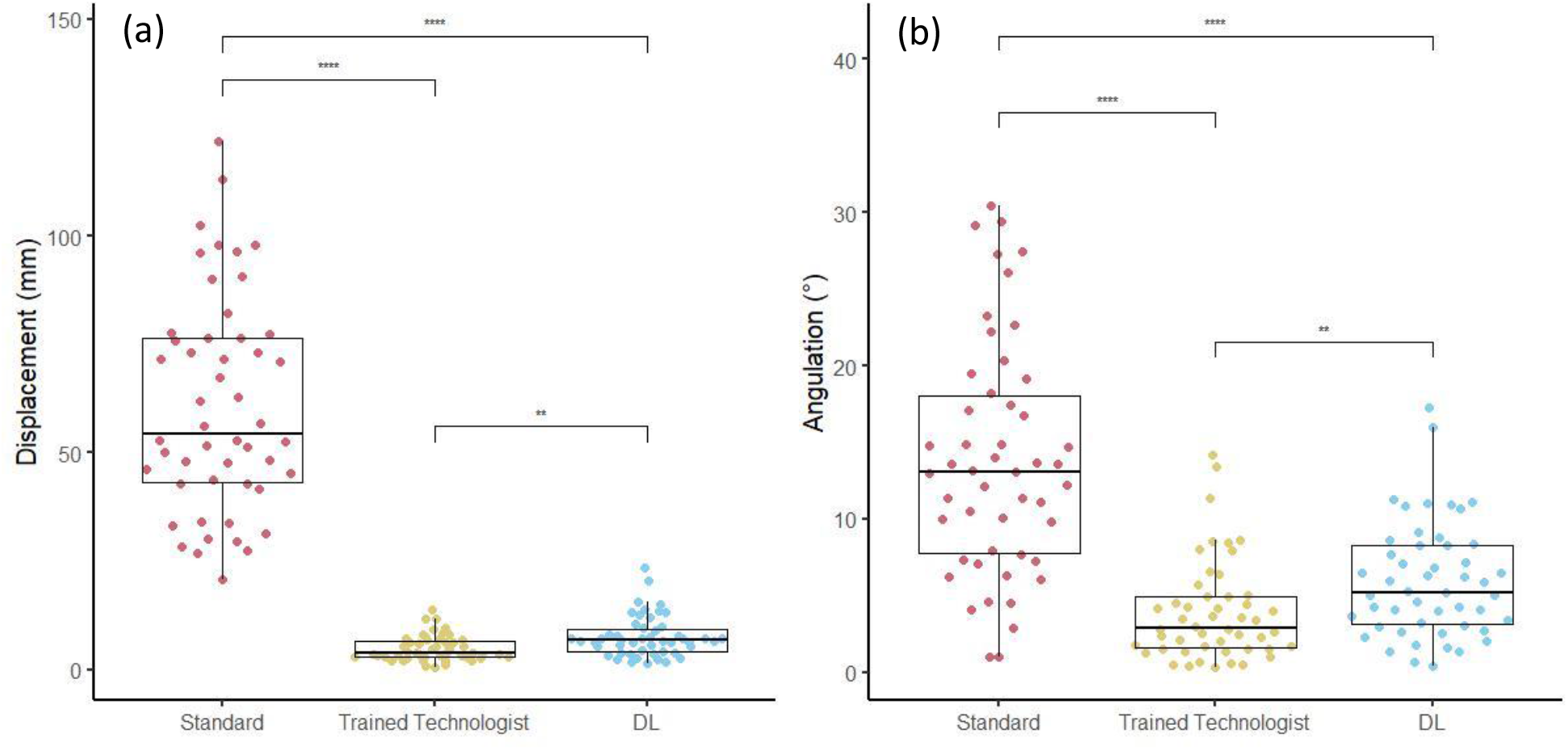
Comparison of the displacement (a) and angulation (b) of the LV for standard, trained user and automatic DL segmentation, versus our expert user segmentation. **: p < 0.01, ***: p < 0.001, ****: p < 0.0001.

The maximum SUV sampled within the LV contours was compared between the imaging expert and the three other processing (trained technologist manual processing, and both automated segmentations) in Figure 4. The DL algorithm showed excellent correlation with the imaging expert (R^2^ = 0.97, p<0.0001), similarly to the trained technologist (R^2^ = 0.98, p<0.0001), and markedly better than the standard segmentation (R^2^ = 0.14, p=0.007).

**Figure 4.**
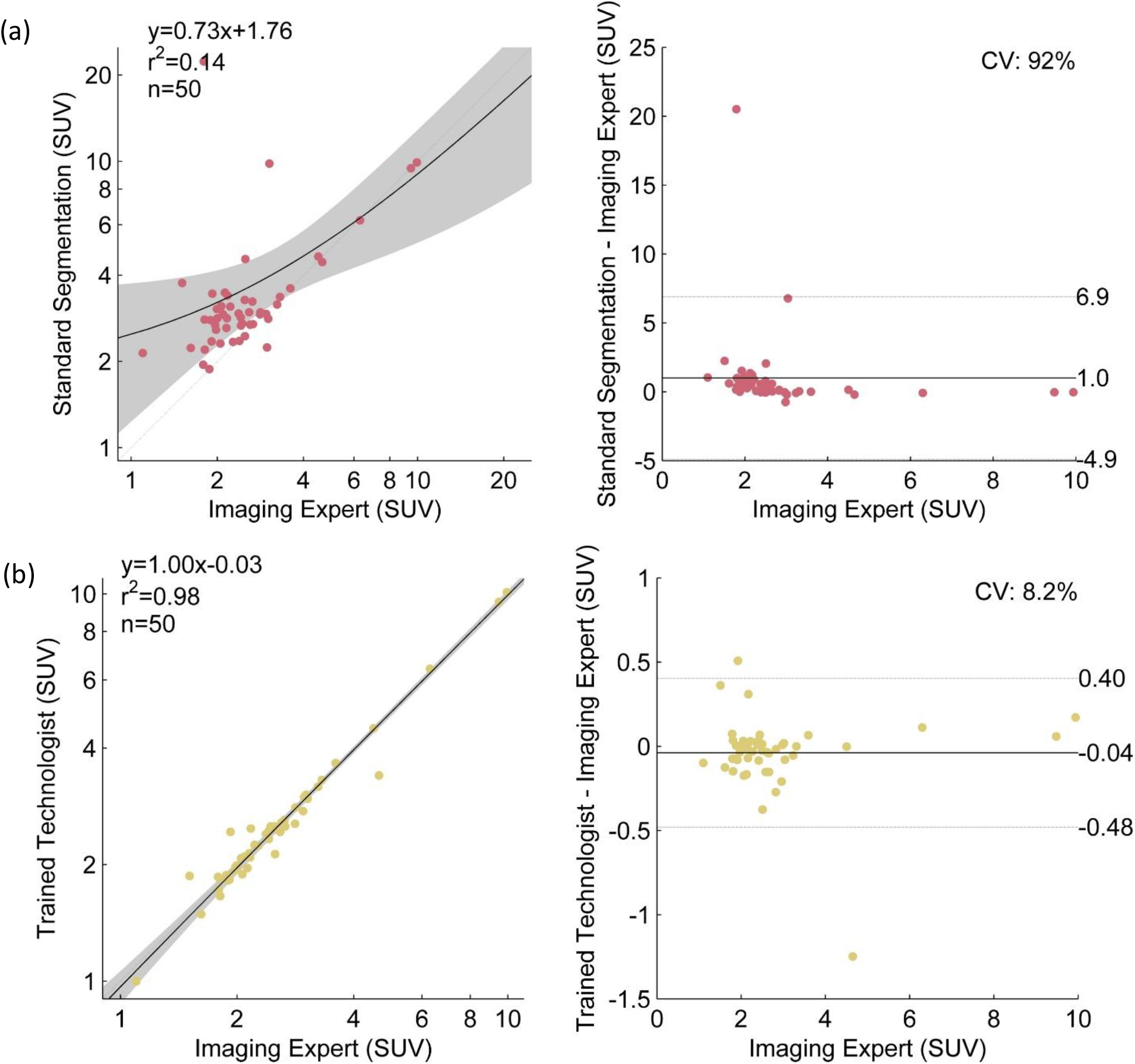
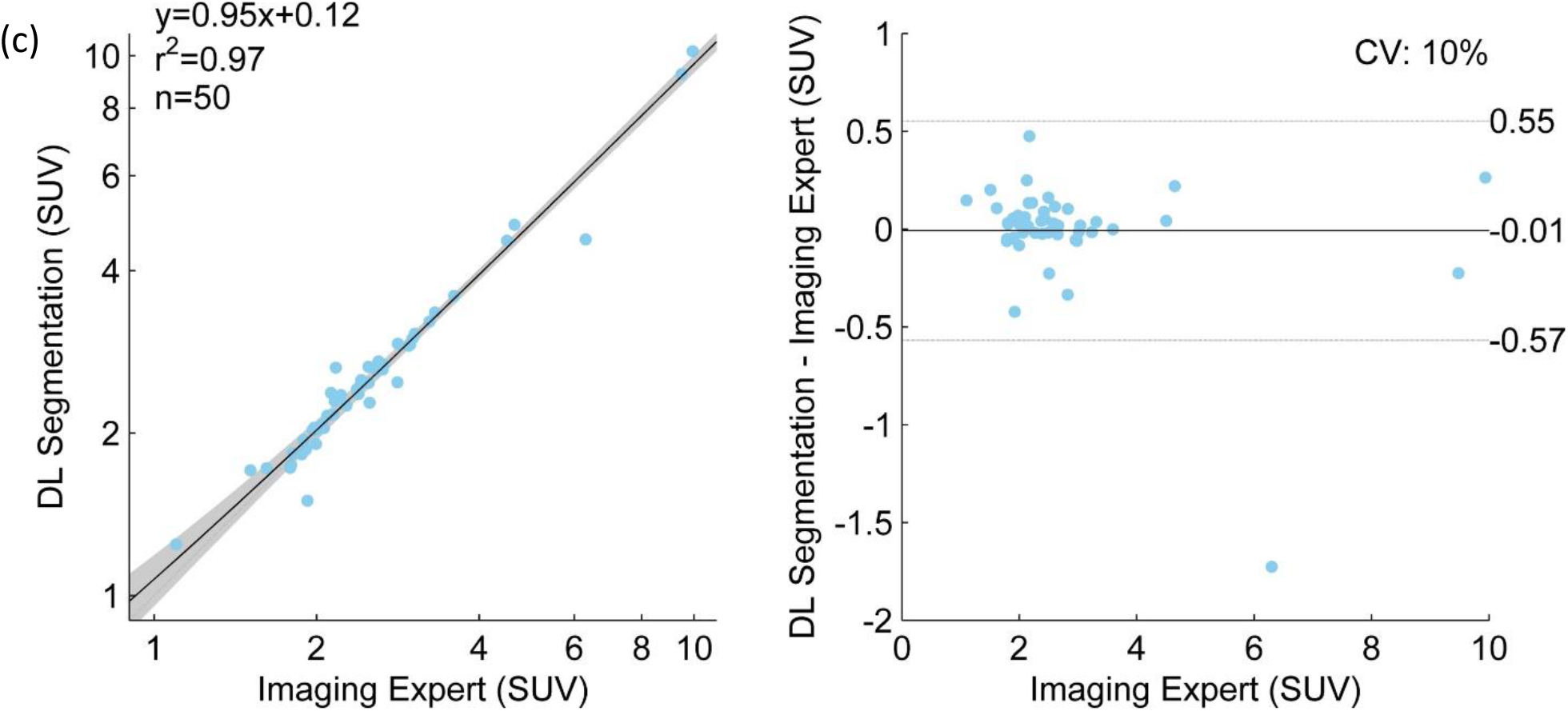
Correlations between our expert user and standard segmentation (a), our trained user (b), and our newly developed DL segmentation (c).

## Discussion

We have developed an automated deep learning algorithm for segmentation of myocardial FDG PET images acquired with a glycolytic metabolic suppression protocol, demonstrating performance comparable to manual contour transfer by a trained technologist. This algorithm markedly improved quality scores assigned by a physician as compared to standard segmentation. This automated algorithm has the potential to improve throughput and increase reproducibility of quantitative and qualitative interpretations.

The scope of this work is focused on image segmentation, reorientation, and scaling for clinical review. Another earlier study has investigated the application of deep learning for the segmentation and classification of FDG PET studies [17]. The focus of that work was on segmentation of a broad heart region including multiple structures rather than the myocardium and on classification performance into no uptake, diffuse uptake, or partial uptake, rather than on optimization of images for clinician review.

While the results show clear performance improvements, there are limitations worthy of consideration for future improvements. This study was performed using data from a single center which has published extensively on optimization of myocardial glycolytic metabolic suppression protocols with generally high-quality studies [5, 6, 13]. As such, implementation at other sites with differing protocols or lower image-quality may not fare as well. Deep learning approaches are rapidly advancing, and U-Net is well established but other networks may have superior performance. Finally, the population used for both construction and validation of the DL model is relatively small and may not contain representative examples of less common cardiac imaging findings.

Ultimately, the latter two limitations suggest further improved results may be feasible using novel deep learning network architectures and larger training and testing databases, including scans from multiple sites with more diversity of protocols and cameras. Having a larger training database is particularly desirable given the diverse activity distributions seen in inflammatory scans, ranging anywhere from multiple high-activity hot spots to no myocardial activity.

Nonetheless, implementation of this new tool within the clinical sarcoid workflow should be a clear success compared to standard segmentation. Cases of inadequate contouring could still be corrected by manual approaches such as transferring contours from perfusion studies, when available. Furthermore, mutual registration of perfusion and FDG datasets will also be more straightforward with improved segmentation of the LV on both image sets.

## Conclusion

In this work, we used deep learning to develop a tool to perform automatic segmentation of cardiac FDG PET studies performed with a myocardial glycolytic metabolic suppression protocol. Results show clear improvement in segmentation, centering, orientation, and normalization resulting in a marked improvement in overall readability of these scans despite the complex and highly variable activity distributions observed. Clinical implementation of this new tool should greatly improve clinical workflow.

## Data Availability

The data used in this study are the property of the University of Michigan and cannot be shared without a data use agreement with them.

## Acknowledgments

The authors acknowledge the Regents of the University of Michigan for the use of de-identified clinical data for this study. Large language models were used to generate the preliminary abstract and introduction of this article, which were then revised by the authors.

